# Inter-diagnostician Agreement and Inter-subject Variability for Binary Diagnostic Decisions in Psychiatry

**DOI:** 10.1101/2020.07.31.20166074

**Authors:** Richard Chalon Aiken

## Abstract

Among the many challenges in arriving at a valid psychiatric diagnosis are the variations in biologic, genetic/ epigenetic, psychological, cognitive, and behavioral attributes of the subject population: *inter-subject variability*. Study of nosologic classification in psychiatry has centered primarily on inter-diagnostician variability, reliability, while neglecting inter-subject variation. We present an analysis of the diagnostic process that considers variability not only in the diagnosticians but also the subjects. The conditions for which the percent agreement statistic is the preferred measure of agreement is detailed for both a general case as well as one for which the inter-subject variation is explicitly considered. Specifics are given as to the non-intuitive fact that the smaller the inter-subject variability, the poorer can be the agreement, suggesting that an agreement statistic alone may be misleading depending on the nature of the subject population. Existing agreement statistics do not measure inter-diagnostician reliability but the agreement between two diagnosticians influenced by the inter-subject variation.

## 1. Introduction

It is challenging to find valid objective diagnostic criteria in the field of psychiatry, currently still quite subjective in nature. Early psychiatric classification, such as that of the American Psychiatric Association Diagnostic and Statistical Manual of Mental Disorders Second Edition (DSM-II) Jackson (1970), based classification upon the *best clinical judgement and experience of a committee and its consultants*, not statistics. The motivation was to form a basis for psychiatric research so that the group of studied subjects had a commonality, akin to assuring experimental reproducibility.

However, after the publication of DSM-II, the well-known Rosenhan experiments, Rosenhan (1973), demonstrated convincingly that the psychiatric classifications had a very low reliability. Enhancing reliability has therefore been a major goal in the development of the DSM-III and subsequent editions.

About the same time in the early 1970s, the more fundamental issue of the validity of the psychiatric diagnosis was formally recognized, arguably first by Robins and Guze (1970). They proposed a five-step plan to investigate the validity of defined psychiatric disorders: clinical description, laboratory studies, exclusion criteria, follow-up studies, and family studies. This focus on diagnostic validity has only recently received serious attention once again with the National Institute of Mental Health’s Research Domain Criteria (RDoC) project First (2012), its goal being *to develop new ways of classifying mental disorders based on dimensions of observable behavior and neurobiological measures*.

## 2. Overview OF THE Diagnostic Process in Psychiatry

In the general medical training that all psychiatrists undergo, one develops the expectation that most presenting acute symptoms can be explained by one diagnosable disease. This is not the case in general psychiatry where related co-morbidities are quite common. Fundamentally, psychiatry is still a subjective science to date and does not focus on actual disease states, distinct process in the body with specific biologic causes and characteristic symptoms. Instead, focus is on irregularity, disturbance, or interruption of normal functions, termed *disorders*; note the title Diagnostic and Statistical Manual of *Mental Disorders*.

Psychiatric science is beginning to recognize that there are psychiatric diseases associated with neurovascular disease, neuroinflammation, oxidative stress, the hypothalamic–pituitary–adrenal axis, the gut-brain association, and neurogenesis, to name a few. It is therefore quite difficult to represent a valid psychiatric diagnosis of a disorder given the multitude of potential biological influences.

Nevertheless, given a description of a psychiatric disorder, valid or not, a diagnostician, whom we shall refer to as a *rater*, meets with an individual, whom we shall call a *subject*, and decides whether or not that subject has the disorder or not based on the given diagnostic criteria. The focus then is whether or not the raters apply the criteria similarly, so-called *inter-rater reliability*.

However, a factor that can profoundly affect the inter-rater reliability is that which confounds the establishment of valid diagnostic criteria as indicated above: *inter-subject variability*.

### 2.1 Inter-subject variation

The challenges of developing valid diagnostic criteria as well as the reliable application of it rest on the inevitable variation of complex and interacting differences amongst the subject population - not only various etiologic biological disease processes as mentioned earlier, but genetic/ epigenetic, psychological, historical, cognitive, and behavioral differences. All of these factors taken together constitute inter-subject variability.

### 2.2 Two raters making a binary diagnostic decision

First, let’s define terms that we shall use in subsequent analyses.

Suppose that two diagnosticians, or raters, use a particular diagnostic method to arrive at a diagnosis of *yes disorder is present*, or *no disorder is present* in a set of subjects, given:

- two raters,
- a set of subjects,
- a diagnostic method.

Consider an *agreement matrix* defined as:

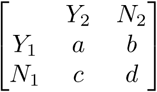

where,

Y1 and N1 = rater 1 choosing disordered and not disordered, respectively,

Y2 and N2 = rater 2 choosing disordered and not disordered, respectively,

a = subjects judged by rater 1 as well as rater 2 to have the disorder,

b = subjects judged by rater 1 as disordered while rater 2 judges the disorder is not present,

c = subjects judged by rater 1 to not have the disorder while rater 2 judges disordered,

d = subjects judged by rater 1 as well as rater 2 to not have the disorder.

Not all of the numbers in these two matrices are independent in that in each matrix, they must sum to the total number of subjects in the study:

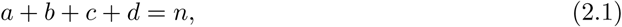

where n is the total number of subjects judged by the two raters in this study. Dividing each element in the agreement matrix by n, gives the *probability matrix* :

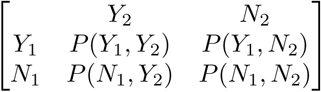

where,

*P* (*Y*_1_, *Y*_2_) = *a/n* is the probability that rater 1 and rater 2 chooses disordered,

*P* (*Y*_1_, *N*_2_) = *b/n* is the probability that rater 1 chooses disordered while rater 2 chooses not disordered,

*P* (*N*_1_, *Y*_2_) = *c/n* is the probability that rater 1 chooses not disordered while rater 2 chooses disordered, and

*P* (*N*_1_, *N*_2_) = *d/n* is the probability that rater 1 and rater 2 chooses not disordered. By definition,

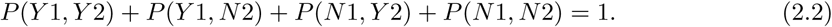

We define the *prevalence*, p, of the disorder as the percentage of a population that is affected with a particular disorder at a given time and assume that in this study that prevalence is represented by the n subjects.

Now we shall turn our attention to mathematically modeling the process of deciding upon a diagnosis in mental health.

### 2.3 Modeling the decision process

Let us define some criterion for making a decision as to whether or not a subject is disordered; let this be a single variable *x* such that if the subject is deemed to have a value of *x* greater than or equal to some fixed value *x*_*c*_, the *criterion value* for a rater, the subject is considered disordered, otherwise the subject is not disordered. In our modeling considerations, it is not necessary that such a variable actually exist, as we shall see, but it might exist.

Let us define the density function *f* (*x*) such that

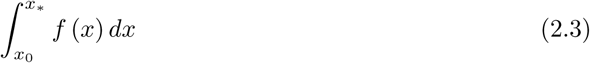

is the probability that the value of *x* is between *x*_0_ and *x*_*\**_.

In particular, we are interested in two probability density functions, one for subjects who are disordered, call that *f*_*y*_(*x*) and another for subjects who are not disordered, call that *f*_*n*_(*x*). These density functions indicate the variation for subjects with respect to the variable *x*. See Figure 1.

**Fig. 1.**
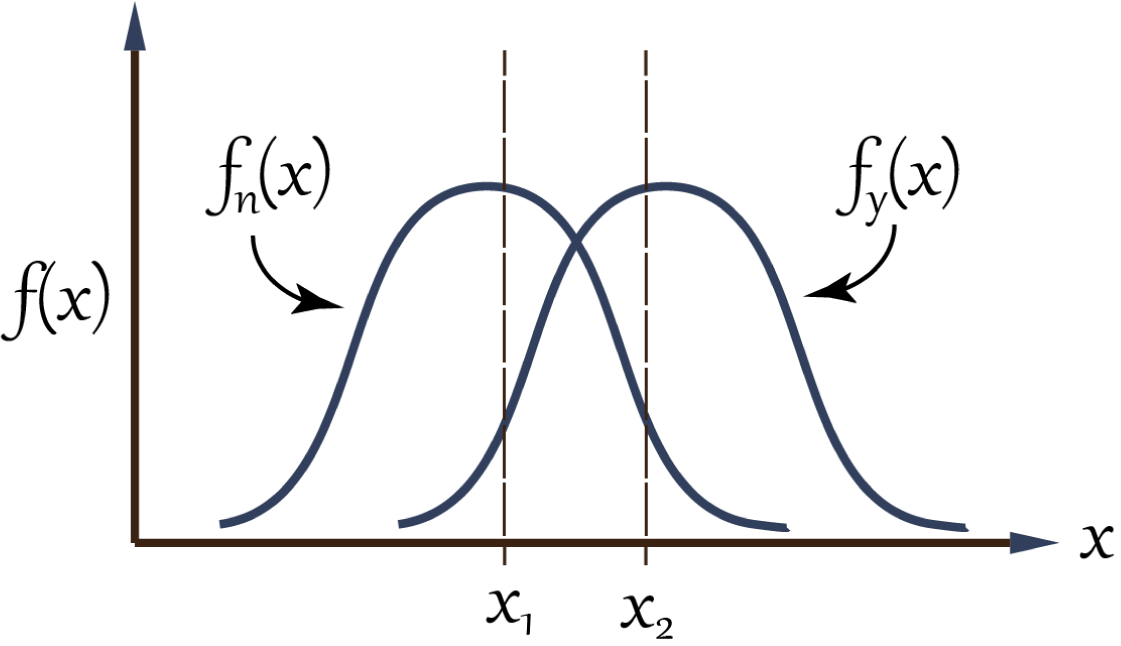
Density functions of those disordered *f*_*y*_ and those not disordered *f*_*n*_ versus the value of *x*, with *x*_1_ and *x*_2_ the values of the criterion variables for rater 1 and rater 2, respectively.

With these functions, we can model the diagnostic method of the two raters in the previous section.

Let *x*_1_ be the criterion value for rater 1 and *x*_2_ be the criterion value for rater 2 as shown in Figure 1.

We shall assume that rater 1 is less conservative about making the disordered diagnosis and that each rater is consistent in their degree of conservatism, or stated differently, there is no *intra-rater dispersion*.

It will be our objective to discover a technique or expression for characterizing the agreement between these two raters.

### 2.4 Inter-rater agreement

Examining the probability matrix, if this simple model is correct, *P*(*N*_1_, *Y*_2_) would always be zero; viz., if there is really a single criterion variable for each rater and by definition *x*_1_ is less than the value of *x*_2_, then it cannot happen that the first rater rates a subject as not disordered and the second rater rates the subject as disordered. The degree to which this matrix element is nonzero is therefore related (staying within the confines of this model) to intra-rater dispersion which, again, we assume for simplicity is negligible.

By the same reasoning, *P*(*Y*_1_, *N*_2_) should be the disagreement between the two raters as rater 1 states disordered while rater 2 states not disordered, the area of the density functions between *x*_1_ and *x*_2_ in Figure 1.

Therefore a teleological measure of agreement between the two raters, let’s call it *ω*, is then

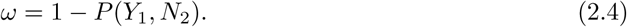

This is known as the *percent agreement statistic*.

Note the conditions for which it is an excellent measure of agreement:

- no intra-rater dispersion,
- each rater maintains a consistent degree of conservatism when making a decision.

Further note that it is not necessary to know:

- the identification of *x*,
- what the criterion variable is for either rater,
- the form of the density functions for subject variability.

The most commonly used agreement measure in practice, intended to represent inter-rater reliability is that of Cohen’s Kappa, Cohen (1960). Cohen used the percent agreement statistic but “corrected” for chance agreement, that is, what agreement would result from random guesses from the raters. In our model, the raters are professionals not making random guesses but systematically and implicitly using a criterion for their choices. We expect that to be much more likely in clinical practice of psychiatry than random guesses. Therefore, correction for random guesses may distort the actual agreement.

## 3. Modeling Pychiatric Decisions

### 3.1 The continuous uniform distribution function

Let’s go back to the fundamental diagnostic process raters use to make a decision. For the binary choice, the diagnostic method consists of each rater setting a decision criterion and applying it to a group of subjects that vary with respect to the variable *x*.

From our previous definitions and equation 2.3,

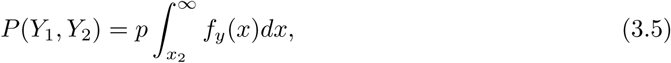

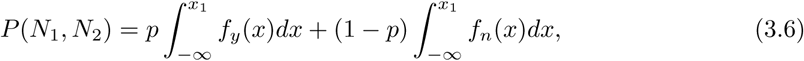

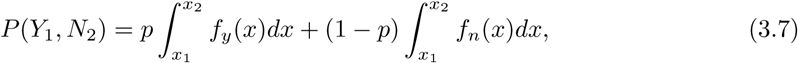

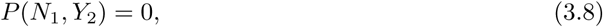

where *f*_*y*_(*x*) is the distribution function of subjects who are actually disordered and *f*_*n*_(*x*) is the distribution function of those subjects who are actually not disordered.

We would like to understand the effect on rater agreement from the selection of *x*_1_ and *x*_2_ for an explicit model of the decision process.

For simplicity we select as our probability density function the continuous uniform distribution:

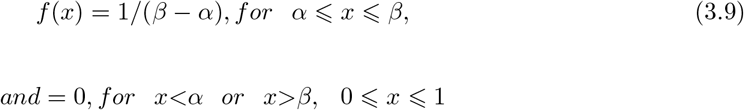

For this model of the subject variability,

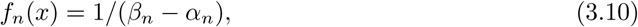

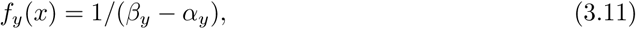

for the bounds given above, where

*α*_*n*_ = the value of *x* at the beginning of the interval for the continuous function *f*_*n*_,

*β*_*n*_ = the value of *x* at the end of the interval for the continuous function *f*_*n*_,

*α*_*y*_ = the value of *x* at the beginning of the interval for the continuous function *f*_*y*_,

*β*_*y*_ = the value of *x* at the end of the interval for the continuous function *f*_*y*_. Integration of equation 3.7 gives,

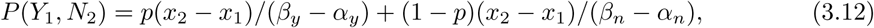

so the agreement statistic is

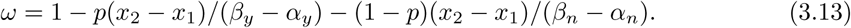

This indicates, in a precise manner, that the larger the inter-subject variability, namely the larger the differences (*β*_*y*_ -*α*_*y*_) or (*β*_*n*_ -*α*_*n*_) or both, the *better* agreement between raters. Therefore, for any given diagnostic method and rater criterion that the effect of inter-subject variability on inter-rater agreement can be quite significant, regardless of the validity of the diagnostic criteria. This has important implications on the interpretation of an agreement statistic. While we wish to measure purely inter-rater reliability we are actually measuring a combination of rater agreement and the variability of subjects with and without the disorder.

Of course, the smaller the inter-rater variability, namely (*x*_2_ − *x*_1_), the better the agreement as well. In fact, in the limit as the differences (*β*_*y*_ − *α*_*y*_) and (*β*_*n*_ − *α*_*n*_) approach unity, indicating complete randomness of characterization of subjects with and without the disorder, 3.12 becomes

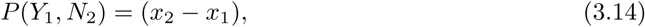

so that,

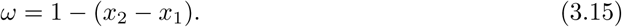

This is the percent agreement statistic of equation 2.4. The interpretation now is that it is a true representation of inter-rater agreement only if the characterization of the subject population is random, otherwise it should properly be termed an agreement statistic.

### 3.2 Small Prevalence and specificity

As the prevalence becomes quite small, a typical case for a sample of the general population, the agreement will depend on the variability of those subjects without the disorder. From 3.13,

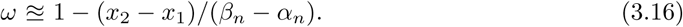

Therefore, the larger the inter-subject variation of those without the disorder, the better the agreement. Equation 3.16 also results for cases in which the variations of the subjects with and without the disorder are equal, regardless of the prevalence.

With very small prevalence, the focus is on specificity, a measure of the proportion of actual negatives that are identified by the two raters. Equation 3.16 indicates that the broader the characteristic of those without the disorder, *β*_*n*_ − *α*_*n*_, the better would be the rater agreement.

### 3.3 Large Prevalence and sensitivity

As the prevalence becomes large, the agreement will depend more on the variability of those subjects with the disorder. From 3.13,

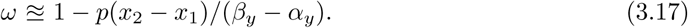

With large prevalence, the focus is on sensitivity, a measure of the proportion of actual positives that are identified by the two raters. Equation 3.17 indicates that the broader the characteristic of those with the disorder, *β*_*y*_ − *α*_*y*_, the better would be the rater agreement.

## 4. Conclusions

Many complex etiologic and individual constitutive factors complicate the establishment of a valid psychiatric classification system. However, this inter-subject variability must be considered in the development, research, and application of any proposed classification system, particularly a subjective one such as the Diagnostic and Statistical Manual series.

Once diagnostic criteria is established, valid or not, it is of interest to check the reliability of its application. Typically this is done by comparing the decisions of raters using an agreement statistic such as Cohen’s Kappa that “corrects” simple agreement for chance agreement. However, in psychiatry and psychology the raters are trained clinicians using explicit diagnostic criteria so that, unlike in some fields of subjective measurement, decisions are much less likely to be mere guesses. This is the reason why we favor a simple agreement statistic.

Just as inter-subject variability confounds the establishment of valid diagnostic criteria, it also can profoundly influence the rater agreement. We have shown, for a specific mathematical representation of the variability in subjects with and without a disorder, that less defined characterization of these populations can actually lead to better, but fortuitous, rater agreement. True inter-rater reliability is not what is measured, rather just rater agreement. Only in the case that the characterization of the subjects is random does it not influence determination of interrater reliability; in that unrealistic case, the percent agreement statistic is a proper inter-rater reliability.

Small prevalence of the disorder leads to agreement that is dependent on the variation of subjects without the disorder, related to specificity; conversely, larger prevalences leads to agreement that is dependent on the variation of subjects with the disorder, related to sensitivity.

While the Diagnostic and Statistical manual is not based on statistics, the statistical analysis given here for the subject population could elucidate the measure of diagnostician agreement.

## Data Availability

no data used

